# Improvements and challenges of long-term survival in patients with systemic lupus erythematosus-associated pulmonary arterial hypertension: A 10-year multi-center cohort study

**DOI:** 10.1101/2023.12.05.23299536

**Authors:** Xingbei Dong, Jiuliang Zhao, Junyan Qian, Wei Wei, Miaojia Zhang, Xiao Zhang, Xiaofei Shi, Yisha Li, Xiaoping Hong, Qiang Shu, Shuhong Chi, Xin Dong, Ping Zhu, Rong Zhang, Zhuoli Zhang, Hongfeng Zhang, Xinwang Duan, Jing Xue, Shuhong Zhou, Hongbin Li, Dan Chen, Junwei Zhang, Yanhong Wang, Zhuang Tian, Yongtai Liu, Mengtao Li, Xiaofeng Zeng, Qian Wang

**Affiliations:** Department of Rheumatology and Clinical Immunology, Peking Union Medical College Hospital, Chinese Academy of Medical Sciences, Peking Union Medical College.; National Clinical Research Center for Dermatologic and Immunologic Diseases (NCRC-DID), Ministry of Science & Technology.; State Key Laboratory of Complex Severe and Rare Diseases, Peking Union Medical College Hospital.; Key Laboratory of Rheumatology and Clinical Immunology, Ministry of Education, Beijing 100730, China; Department of Rheumatology, Tianjin Medical University General Hospital, Tianjin, China; Department of Rheumatology, the First Affiliated Hospital of Nanjing Medical University, Nanjing, China; Department of Rheumatology, Guangdong General Hospital, Guangzhou, China; Department of Rheumatology, the First Affiliated Hospital of Henan University of science and Technology, Luoyang, China; Department of Rheumatology, Xiangya Hospital, Central South University, Changsha, China; Department of Rheumatology, Shenzhen People’s Hospital, Shenzhen, China; Department of Rheumatology, Qilu Hospital of Shandong University, Jinan, China; Department of Rheumatology, General Hospital of Ningxia Medical University, Yinchuan, China; Department of Rheumatology, Beijing Chao-Yang Hospital, Capital Medical University, Beijing, China; Department of Clinical Immunology, PLA Specialised Research Institute of Rheumatology and Immunology, Xijing Hospital, Fourth Military Medical University, Xi’an, China; Department of Rheumatology, the First Hospital of China Medical University, Shenyang, China; Department of Rheumatology and Clinical Immunology, Peking University First Hospital, Beijing, China; Department of Rheumatology, the First Affiliated Hospital of Dalian Medical University, Dalian, China; Department of Rheumatology, the Second Affiliated Hospital of Nanchang University, Nanchang, China; Department of Rheumatology, the Second Affiliated Hospital of Zhejiang University School of Medicine, Hangzhou, China; Department of Rheumatology, Gansu Provincial Hospital, Lanzhou, China; Department of Rheumatology, the Affiliated Hospital of Inner Mongolia Medical University, Hohhot, China; Department of Rheumatology, the First Affiliated Hospital of Wenzhou Medical University, Wenzhou, China; Department of Critical Respiratory Medicine, Beijing Chao-Yang Hospital, Capital Medical University, Beijing, China; Department of Epidemiology and Bio-statistics, Institute of Basic Medical Sciences, Chiness Academy of Medical Sciences and Peking Union Medical College, Beijing, China; Department of Cardiology, Peking Union Medical College Hospital, Peking Union Medical College and Chinese Academy of Medical Sciences, Beijing, China

**Author notes:** Xingbei Dong, Jiuliang Zhao, Junyan Qian, and Wei Wei contributed equally to this study. Correspondence to: Prof Mengtao Li; Prof Xiaofeng Zeng; Prof Qian Wang. Department of Rheumatology and Clinical Immunology, Peking Union Medical College Hospital, Chinese Academy of Medical Sciences, Peking Union Medical College. National Clinical Research Center for Dermatologic and Immunologic Diseases (NCRC-DID), Ministry of Science & Technology. State Key Laboratory of Complex Severe and Rare Diseases, Peking Union Medical College Hospital. Key Laboratory of Rheumatology and Clinical Immunology, Ministry of Education, Beijing 100730, China.

**Keywords:** pulmonary arterial hypertension, systemic lupus erythematosus, long-term survival, lupus low-disease-activity state, dual treat-to-target strategy

## Abstract

**Background:** Prior studies indicated improved survival in systemic sclerosis-associated pulmonary arterial hypertension (PAH) patients, but trends in systemic lupus erythematosus-associated PAH (SLE-PAH) survival remained unclear.

**Methods:** Analysing SLE-PAH patients from the nationwide CSTAR-PAH cohort, we divided them into two cohorts: A (2011-2016) and B (2016-2021), based on baseline right heart catheterization dates. We compared clinical characteristics, mortality, and treatment outcomes between these cohorts and with idiopathic PAH (IPAH) patients.

**Results:** We enrolled 610 and 104 patients with SLE-PAH and IPAH, respectively. Patients with SLE-PAH were younger, had a higher proportion of low-risk patients, and had a significantly higher 10-year survival rate than those with IPAH (66·6% vs. 44·1%, p < 0·001). Cohort B had a longer 6-min walk distance, lower mean pulmonary arterial pressure and pulmonary vascular resistance, a better-preserved cardiac index, and less right ventricular dilation than cohort A. More patients in cohort B received intensive immunosuppressant- and PAH-targeted therapies. The 5-year survival rate was significantly higher in cohort B (88·1% vs. 77·5%, p = 0·006). Reaching low-risk profile of PAH (hazard ratio [HR] 0·34, 95% confidence interval [CI] 0·15-0·79, p = 0·012) and reaching lupus low-disease-activity state (HR 0·33, 95% CI 0·14-0·82, p = 0·016) were independent predictors of survival. The rate of achieving low-risk profile for PAH was considerably higher in patients initially treated with intensive immunosuppressive and dual-PAH-targeted therapies.

**Conclusions:** Over the last decade in China, the clinical characteristics of patients with SLE-PAH have evolved and survival has improved. Early PAH detection and dual treatment-to-target strategies for both PAH and SLE have contributed to this improvement in survival.

*What is new?:*

- This is the largest multi-center prospective cohort study of SLE-PAH with the longest follow-up period describing changes in the characteristics, treatment regimen, and long-term survival of patients with SLE-PAH.
- Our study showed that the 5-year survival rate of patients with SLE-PAH has increased remarkably from 77·5% to 88·1% during the last decade.
- Our study demonstrated that reaching lupus low-disease-activity state is independently associated with reduced mortality. Significantly more patients reached low-risk profile of PAH during follow-up with initiation of intensive immunosuppressive therapy.

*What are the clinical implications?:*

- Our study emphasised on the importance of achieving dual treatment goals for both SLE and PAH (dual treat-to-target strategy).
- Earlier detection of PAH in patients with SLE, timely initiation of intensive immunosuppressive therapy, and upfront combination PAH-targeted therapy benefit patients in achieving PAH low-risk profile.

## Introduction

Pulmonary arterial hypertension (PAH) is a pathophysiological disorder characterised by progressive pressure elevation in the pulmonary circulation due to pulmonary vascular disease, eventually leading to right heart failure^1^. Connective tissue disease (CTD) is one of the major aetiologies of PAH, accounting for 13·1%–25·3% of cases of PAH^2–4^. Meanwhile, PAH is a severe and frequent complication of CTD and remains a leading cause of mortality in patients with CTD^5^.

Over the last two decades, the development of PAH medications targeting the endothelin-1, nitric oxide, and prostaglandin I_2_ pathways and the constant refinement of treatment strategies have led to improvements in the clinical outcomes of patients with PAH^3, 4, 6^. A recent study performed at Johns Hopkins Pulmonary Hypertension Center reported that transplant-free survival markedly improved in patients with systemic sclerosis (SSc)-associated PAH (SSc-PAH) ^7^.

Unlike in Western populations, systemic lupus erythematosus (SLE)-associated PAH (SLE-PAH) is the leading cause of CTD-associated PAH (CTD-PAH) in China, accounting for approximately 50% of the cases^8^. SLE-PAH shows very different outcomes from SSc-PAH and other types of pulmonary hypertension^9–11^, probably because of the remarkable inflammatory nature of SLE-PAH. However, whether the evolving diagnostic and treatment strategies benefit patients with SLE-PAH in a real-world setting remains unclear.

Therefore, we conducted this multi-center prospective cohort study to explore changes in the clinical characteristics, initial treatment strategy, treatment goal achievement, and long-term survival of patients with SLE-PAH over the past decade. We also discuss possible reasons for the improvement in survival and describe the challenges that remain in the disease management of SLE-PAH.

### Methods Study design

The Chinese SLE Treatment and Research Group (CSTAR) PAH cohort is a nationwide multi-center prospective cohort established in 2006, involving 21 qualified CTD-associated PAH referral centers (Supplement Table 1) in China^12, 13^. Ethical approval was granted by by the Institutional Review Board of Peking Union Medical College Hospital (PUMCH) (JS-2038). All patients in the cohort met the 1997 American College of Rheumatology classification^14^ or the 2012 Systemic Lupus International Collaborating Clinics classification^15^ for SLE. PAH was defined as a resting mean pulmonary arterial pressure (mPAP) of ≥25 mmHg, pulmonary arterial wedge pressure of ≤15 mmHg, and pulmonary vascular resistance (PVR) of >3 Wood units (WU) obtained by right heart catheterisation (RHC). Patients with SLE-PAH diagnosed between 1 June 2011, and 31 May 2021, were enrolled in the study. Patients with idiopathic PAH (IPAH) were consecutively recruited from Beijing Chaoyang Hospital as the control group during the same period to simultaneously describe the baseline characteristics and survival of patients with SLE-PAH. Patients who did not fulfil the hemodynamic definition of pre-capillary pulmonary hypertension and those associated with conditions such as human immunodeficiency virus infection, portal hypertension, congenital heart disease, heart failure with reduced ejection fraction, severe lung disease, or pulmonary embolism were excluded. Severe lung disease was defined as a total lung capacity <70% of the predicted value or signs of extensive pulmonary fibrosis on high-resolution chest computed tomography^16^. Eligible patients with SLE-PAH were divided into cohorts A and B according to the date of RHC diagnosis. Cohort A was recruited from 1 June 2011, to 31 May 2016, and cohort B underwent RHC from 1 June 2016, to 31 May 2021.

### Data collection

Baseline was defined as the time of RHC diagnosis. The following baseline variables were selected: age; sex; disease duration from diagnosis of SLE; onset symptoms of PAH; laboratory results, including serum chemistry panel and N-terminal pro-brain natriuretic peptide (NT-proBNP) level; World Health Organization function class (WHO-FC); 6-minute walk distance (6-MWD); hemodynamic parameters; transthoracic echocardiogram (TTE) findings; pulmonary function test (PFT); SLE Disease Activity Index 2000 (SLEDAI-2K) ^17^; and initial treatment regimens. Patients were also stratified according to their baseline characteristics using variables selected from the risk assessment strategy proposed in the 2022 ECS/ERS Guidelines^18^ (the parameters are shown in Supplement Table 2). The low-, intermediate-, and high-risk groups were assigned scores of 1, 2, and 3, respectively. The average score was then calculated for each patient and rounded to the nearest integer. The primary endpoint was the all-cause mortality rate. The secondary endpoint was reaching low-risk profile of PAH according to the four-stratum risk assessment model COMPERA 2.0^19^. Moreover, follow-up data were collected at each clinic visit to determine the achievement of lupus low-disease-activity state (LLDAS) ^20^, which was determined if (1) SLE-DAI-2K ≤4, with no activity in major organ systems; (2) no new lupus disease activity compared with the last assessment; (3) physician global assessment score ≤1; and (4) prednisolone (or equivalent) dose ≤7.5 mg daily with well-tolerated maintenance doses of immunosuppressants.

### Statistical analysis

Mean and standard deviation were used to describe the parametric data. The independent *t*-test, analysis of variance (ANOVA), and Kruskal–Wallis *H* test were used to compare continuous variables. The chi-squared test or Fisher’s exact test was used to compare categorical variables. Statistical significance was set at p <0·05. Survival analysis was performed using the Kaplan–Meier method, and the log-rank test was used to compare survival rates. Univariate and multivariate Cox regression analyses were used to identify factors associated with all-cause mortality. Because reaching low-risk profile of PAH and reaching LLDAS of SLE were time-dependent covariates, we applied time-dependent Cox regression to avoid immortal time bias. Also, additional survival analyses using propensity score matching for age, sex, and cardiac index (CI) were performed, and the proportion of patients reaching a low-risk profile of PAH during follow-up was compared between matched samples with different treatment regimens. SPSS version 26 (SPSS, Chicago, IL, USA) was used for all statistical analyses.

## Results

### Baseline clinical characteristics of patients with SLE-PAH and IPAH

In total, 610 eligible patients with SLE-PAH were included in this study. Among these, 314 and 296 patients were included in cohorts A and B, respectively (Figure 1). In total, 104 eligible patients with IPAH were recruited for this study. Eighteen patients with SLE-PAH (3·0%) were lost to follow-up and were excluded from the survival analysis.

**Figure 1.**
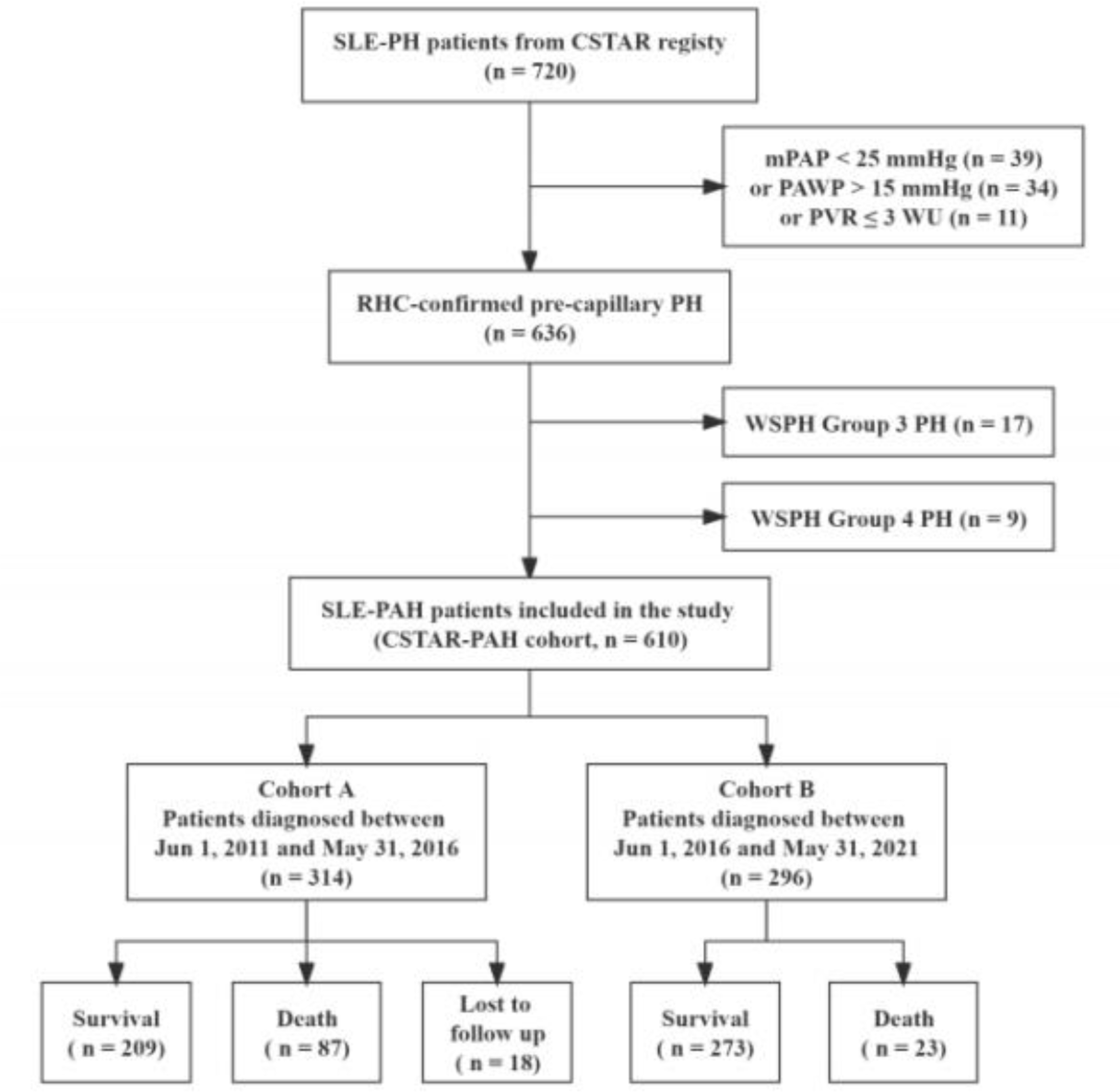
Study flow diagram. SLE, systematic lupus erythematosus; PH, pulmonary hypertension; CSTAR, Chinese SLE Treatment and Research Group; mPAP, mean pulmonary arterial pressure; PAWP, pulmonary arterial wedge pressure; PVR, pulmonary vascular resistance; WU, wood units; RHC, right heart catheterization; WSPH, World Symposium on Pulmonary Hypertension; PAH, pulmonary arterial hypertension.

Comparisons of the demographic and clinical characteristics between the SLE-PAH and IPAH groups as well as between SLE-PAH cohorts A and B are shown in Table 1. Compared with patients with IPAH, patients with SLE-PAH were younger (35·2 ± 9·9 vs. 41·8 ± 16·2 years, p < 0·001) and more predominantly female (98·5% vs. 69·2%, p < 0·001). Pulmonary hypertension in patients with SLE-PAH was less severe than that in patients with IPAH, with fewer patients presenting with manifestations such as exertional dyspnoea and syncope, better exercise capacity, lower NT-proBNP level, less right ventricular dilation or reduced right ventricular function on echocardiography, and a better overall hemodynamic profile.

**Table 1.**
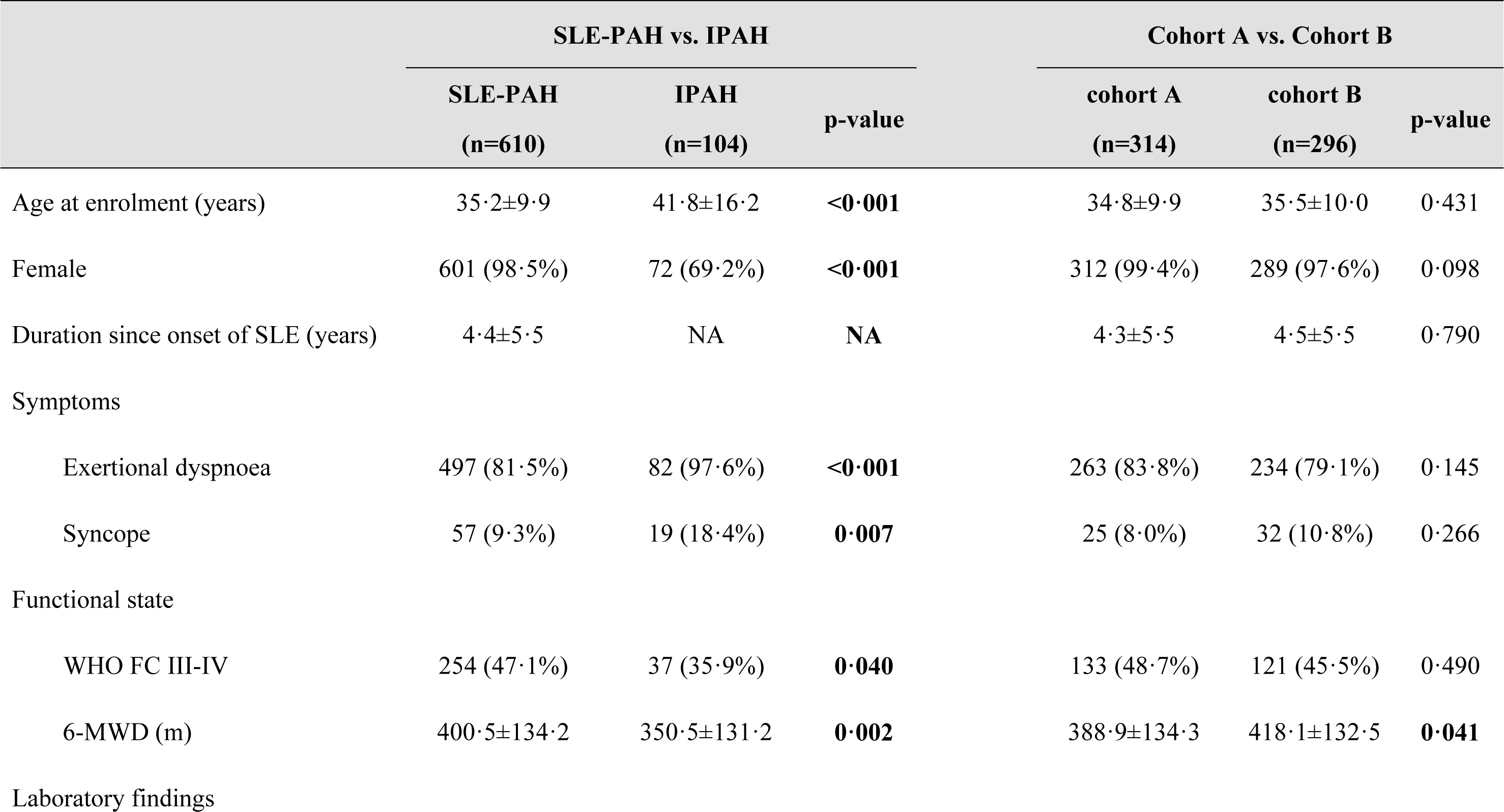
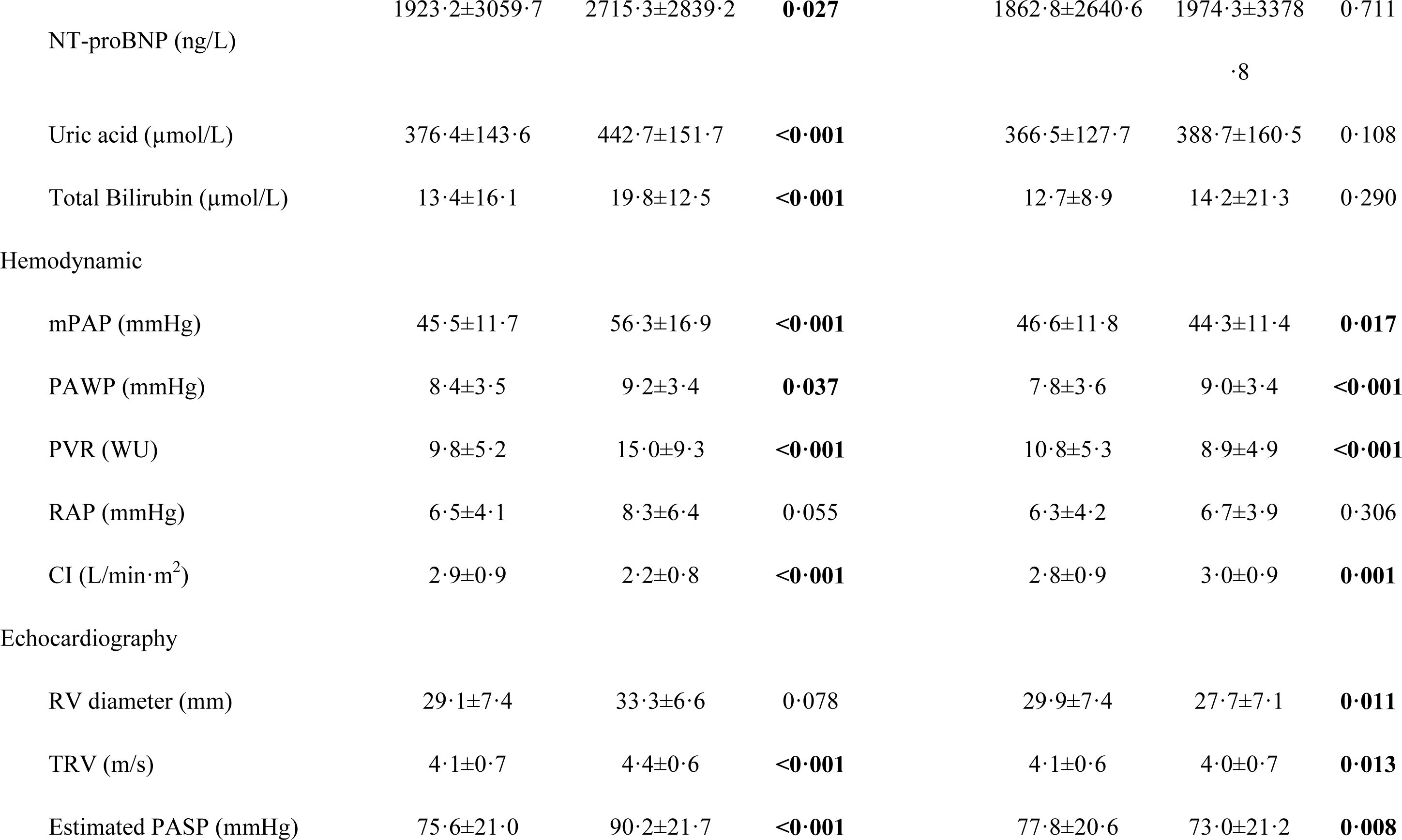
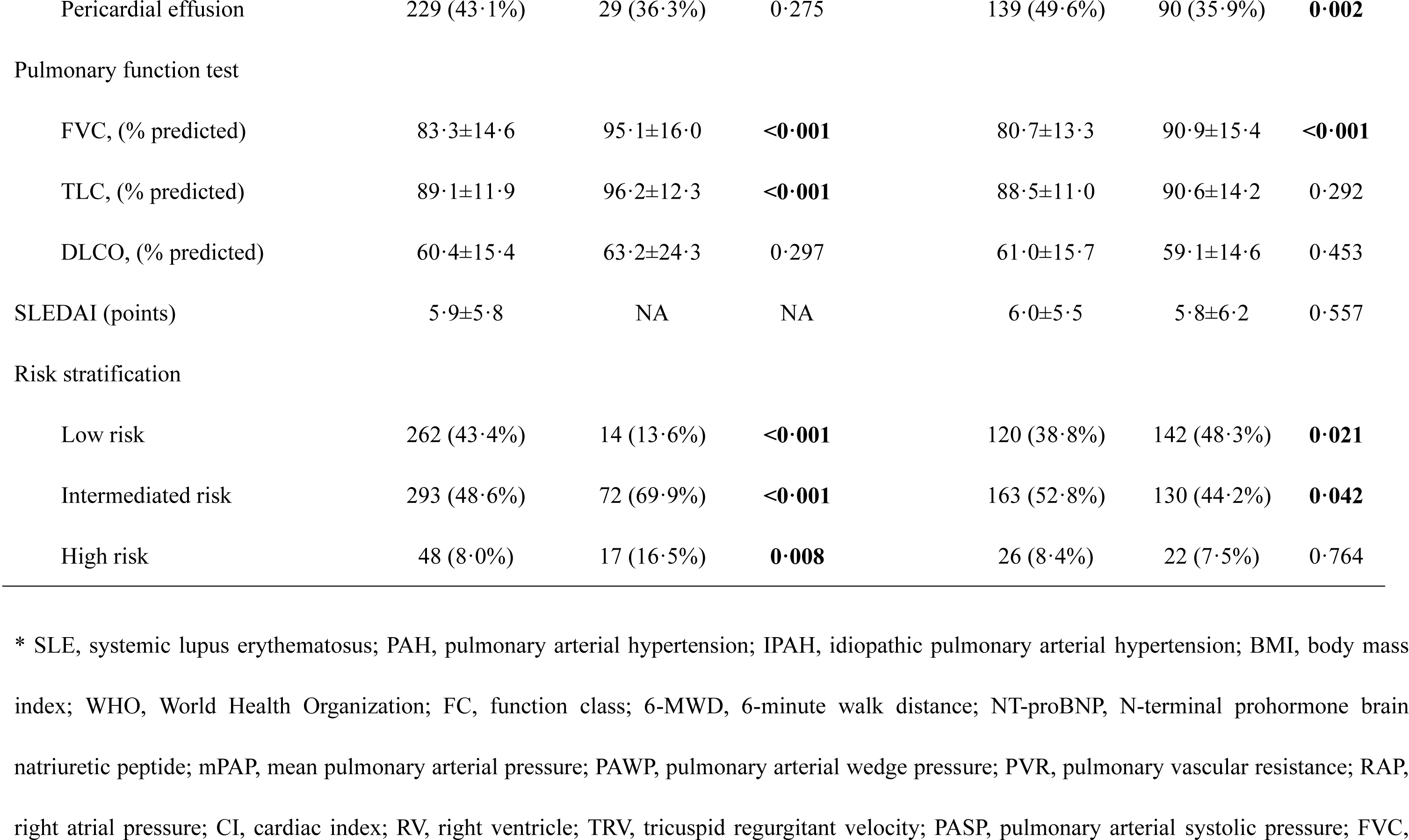
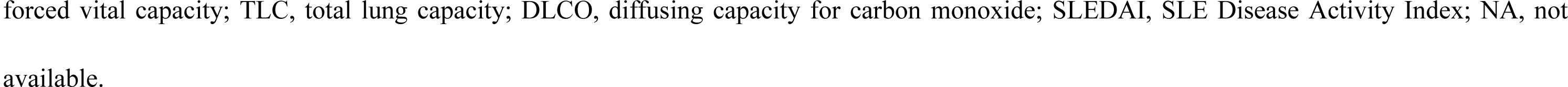
Baseline characteristics of patients with SLE-PAH and patients with IPAH.

The demographic characteristics and SLEDAI were similar between SLE-PAH cohorts A and B. Patients in cohort B showed better performance in terms of 6-MWD (418·1 ± 132·5 vs. 388·9 ± 134·3 m, p = 0·041), a smaller mean right ventricular diameter (27·1 ± 7·1 vs. 29·9 ± 7·4 mm, p = 0·011), and less pericardial effusion (35·9% vs. 49·6%, p = 0·002) on TTE as well as improved hemodynamics with a lower mPAP (44·3 ± 11·4 vs. 46·6 ± 11·8 mmHg, p = 0·017), lower PVR (8·9 ± 4·9 vs. 10·8 ± 5·3 WU, p < 0·001), and preserved CI (3·0 ± 0·9 vs. 2·8 ± 0·9 L/min/m^2^, p = 0·001). More patients in cohort B presented with a low-risk profile than those in cohort A at baseline; however, the percentage of high-risk patients was similar between the two cohorts (Table 1 and Supplement Figure 1).

### Initial treatment regimen of patients with SLE-PAH

As shown in Table 2, most patients with SLE-PAH were treated with glucocorticoids (99·0%) or immunosuppressants (93·6%). Compared to cohort A, more patients in cohort B were treated with intensive immunosuppressive therapy (IIT) (79·7% vs 67·0%, p < 0·001), defined as one or more immunosuppressants, including cyclophosphamide (CYC), calcineurin inhibitors (CNIs), and mycophenolate mofetil (MMF). More patients in cohort B were treated with PAH-targeted therapy (90·9% vs. 67·2%, p < 0·001) and dual PAH-targeted therapy (41·9% vs. 12·7%, p < 0·001). We then compared the initial treatment regimen between cohort A and cohort B in different risk groups, and showed the significant difference in low to intermediate group (Supplement Table 3). There was no significant difference in the high-risk group between cohort A and B.

**Table 2.**
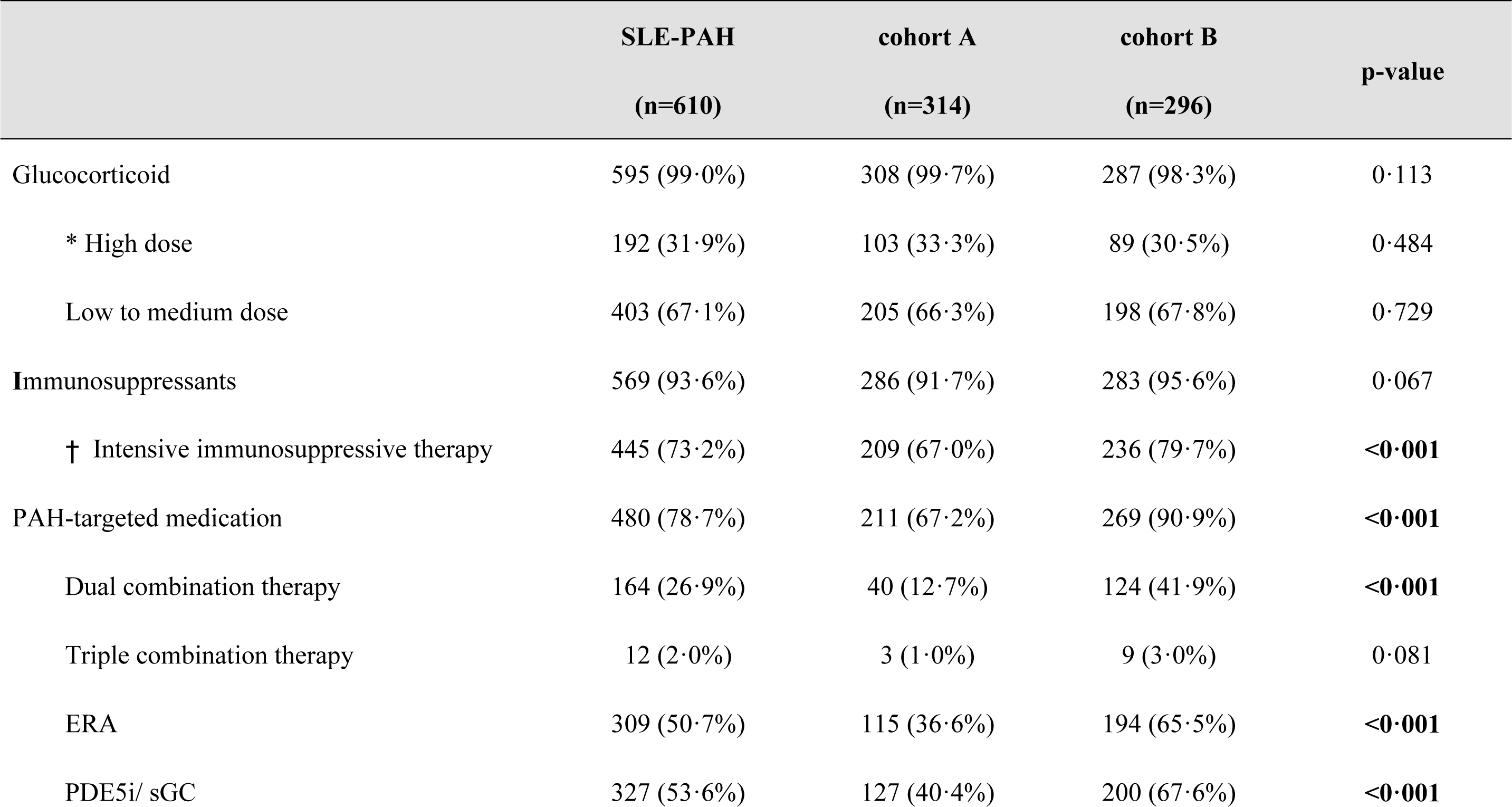
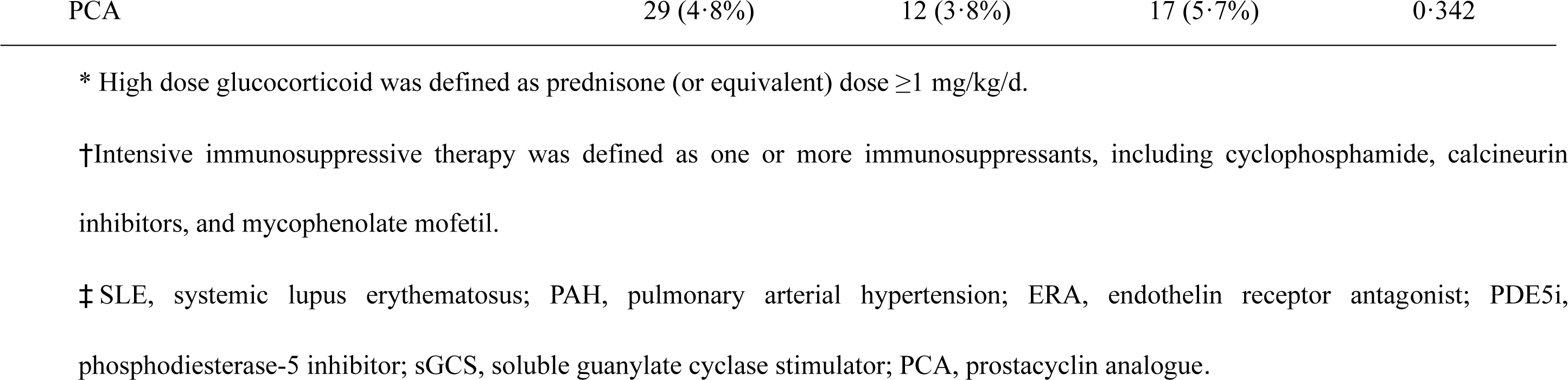
Initial treatment regimen for patients with SLE-PAH.

### Prognosis of patients with SLE-PAH and patients with IPAH

The average follow-up time of patients with SLE-PAH was 4·6 ± 3·0 years. A total of 110 patients with SLE-PAH (18·0%) died during the follow-up. The 5- and 10-year survival rates of SLE-PAH were 81·2% and 66·6%, respectively, which were significantly higher than patient with IPAH (61·7% and 44·1%, respectively, p < 0·001, Figure 2A). Among patients with SLE-PAH, the overall survival rate was significantly higher in cohort B than in cohort A (p = 0·006, Figure 2B). The 1-, 3-, and 5-year survival rates were 93·2%, 85·0%, and 77·5%, respectively, in cohort A and 96·2%, 91·4%, and 88·1%, respectively, in cohort B. When comparing survival between SLE-PAH cohorts A and B stratified into different risk groups, an improvement in survival was observed in the low- and intermediate-risk groups (5-year survival rate 81·7% vs. 89·5%, p = 0·023, Figure 2D) but not in the high-risk group (5-year survival rate 35·4% vs. 69·5%, p = 0·160, Figure 2E).

**Figure 2.**
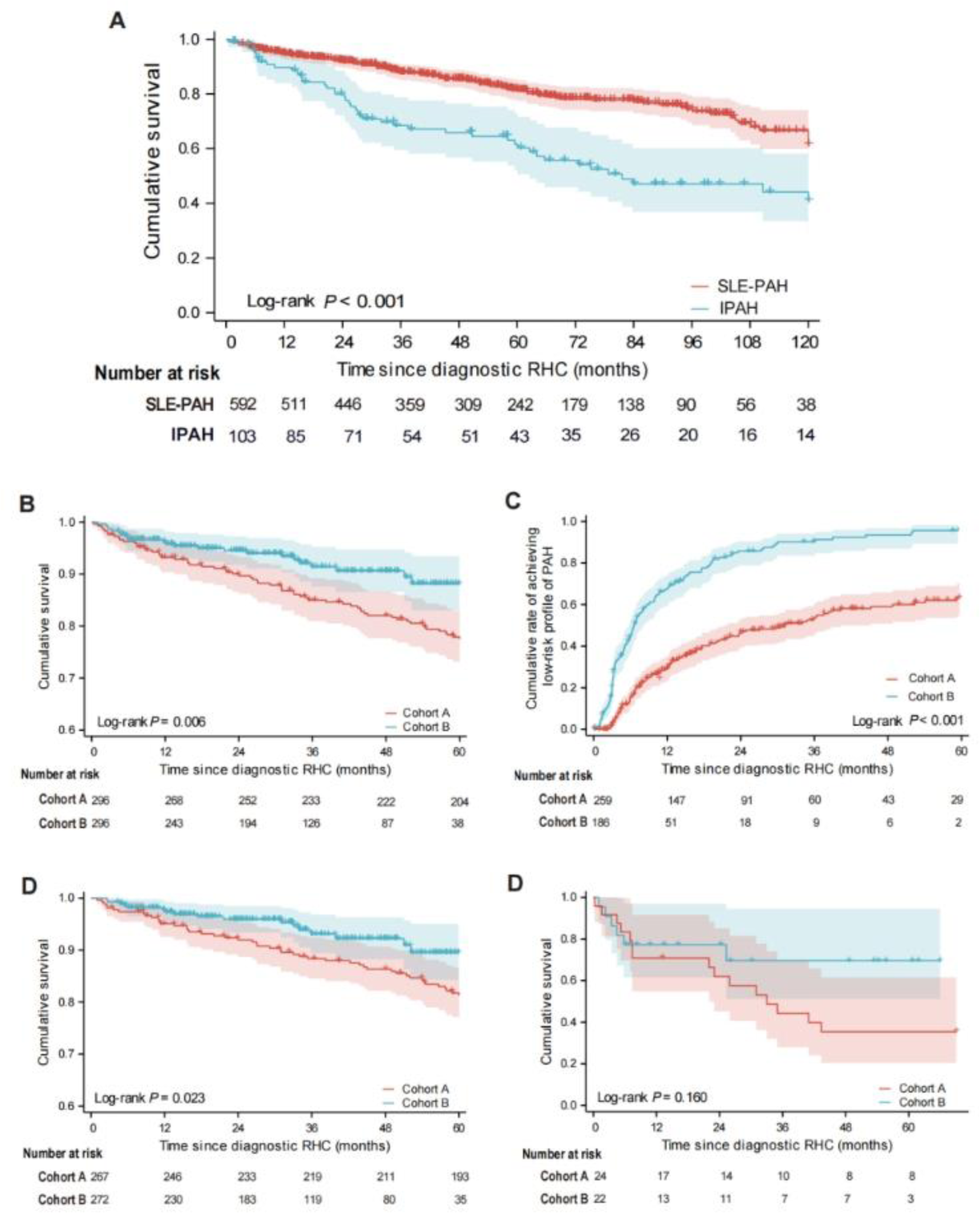
Kaplan–Meier survival analysis. (A) Comparison of 10-year survival between patients with SLE-PAH and patients with IPAH. (B) Comparison of 5-year survival between cohorts A and B. Comparison of 5-year survival between patients with SLE-PAH in (C) Comparison of rate of achieving low-risk profile of PAH during follow-up between cohorts A and B. (D) Comparison of 5-year survival between cohort A and B in low and intermediate-risk groups and (E) in high-risk group. SLE, systemic lupus erythematosus; PAH, pulmonary arterial hypertension; IPAH, idiopathic pulmonary arterial hypertension·

During follow-up, 304 patients with SLE-PAH (66·1%) reached low-risk PAH profile and 287 (82·0%) patients reached LLDAS. Compared with cohort A, more patients in cohort B reached low-risk profile of PAH (Figure 2C, p < 0·001). The median time of reaching LLDAS was significantly shorter in cohort B compared with cohort A (11·5 ± 1·0 vs. 29·7 ± 2·4, p < 0·001).

### Prognostic factors for patients with SLE-PAH

Univariate analysis was used to identify prognostic factors for SLE-PAH. Baseline characteristics, including PAH as an onset symptom of SLE, WHO-FC III–IV, reduced 6-MWD, elevated NT-proBNP concentration, elevated uric acid, elevated total bilirubin, increased PVR, reduced CI, large right ventricular diameter, presence of pericardial effusion, and diffusing capacity of the lung for carbon monoxide of <45% of the predicted value were prognostic factors associated with all-cause mortality (Supplement Table 4).

As shown in Table 3, after adjusting for confounding factors, including age, sex, and CI, reaching a low-risk profile of PAH was independently associated with a reduced risk of death (hazard ratio [HR] 0·34, 95% confidence interval [CI] 0·15-0·79, p = 0·012). LLDAS, a widely accepted treatment goal for SLE, was also favourably associated with better survival (HR 0·33, 95% CI 0·14-0·82, p = 0·016) in muti-variant time-dependent Cox regression.

**Table 3.**
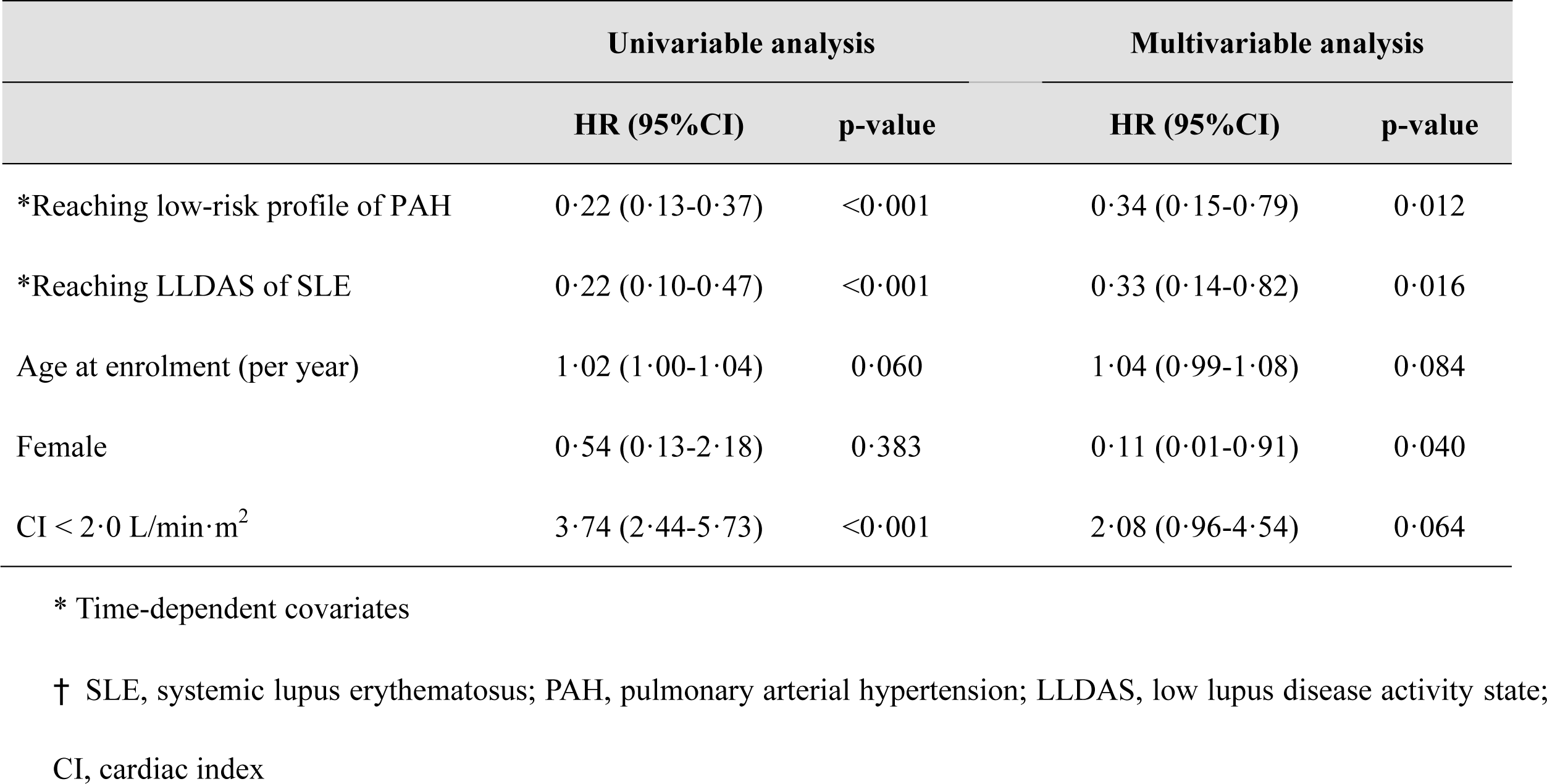
Predictors of all-cause mortality in patients with SLE-PAH.

To further explore the prognostic value of initial treatment regimens in patients with SLE-PAH, we compared the rate of reaching low-risk profile of PAH between propensity score-matched patients with SLE-PAH who were initially treated with and without IIT, as well as between patients who received mono PAH-targeted therapy versus dual PAH-targeted therapy. Notably, patients who were initially treated with IIT (Figure 3A, p < 0·001) and dual PAH-targeted therapy (Figure 3B, p = 0·038) were more likely to reach low-risk profile of PAH during follow-up.

**Figure 3.**
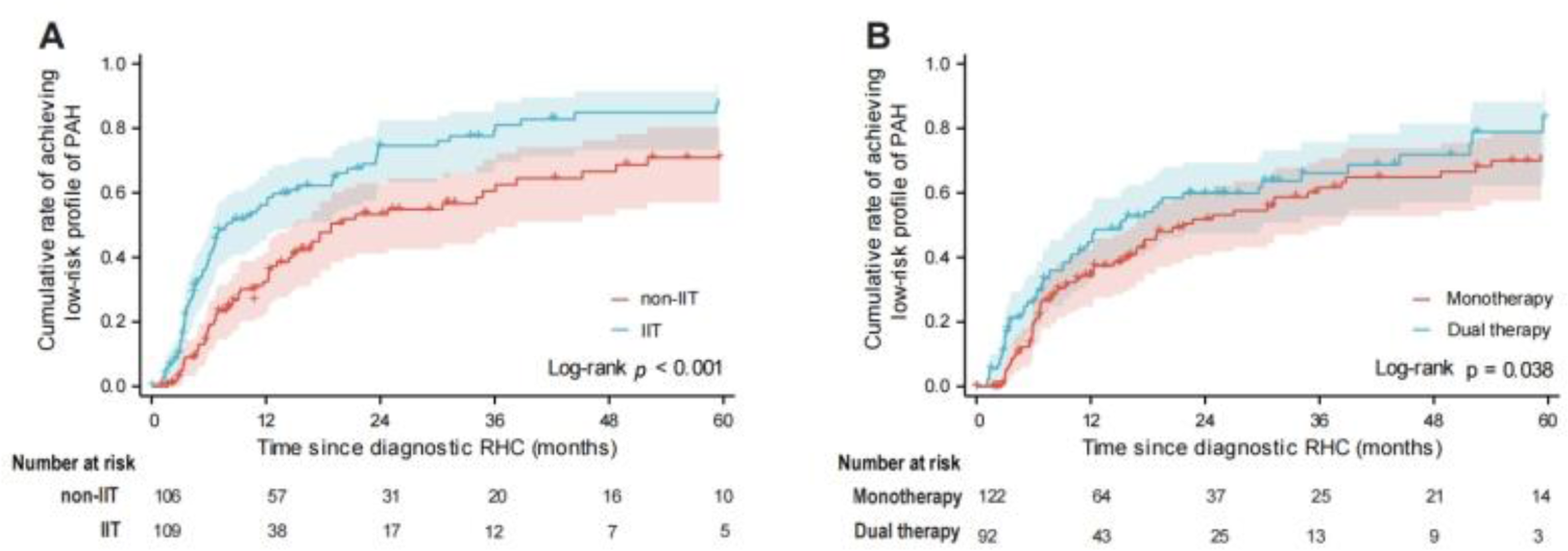
Comparison of rate of achieving low-risk profile of PAH during follow-up between propensity score-matched groups. (A) Comparison with and without IIT and (B) with mono-versus dual PAH-targeted therapy. PAH, pulmonary arterial hypertension; IIT, intensive immunosuppressive therapy.

## Discussion

In this multi-center prospective cohort study of patients with SLE-PAH, we compared changes in baseline characteristics, initial treatment regimens, and overall survival over the last decade. We also identified factors associated with prognosis. We found that more patients were diagnosed at an earlier stage of PAH and were treated with IIT and PAH-targeted therapies in the last five years. Survival rates for SLE-PAH have extensively improved over the past decade. Notably, achieving low-risk profile for PAH and LLDAS for SLE were identified as independent predictors of survival in patients with SLE-PAH. Patients treated with IIT and dual PAH-targeted therapy were more likely to achieve a low risk profile for PAH during follow-up.

Previous studies have suggested that patients with SLE-PAH have much better short-term outcomes than those with IPAH or SSc-PAH. Chung et al.^9^ compared the clinical characteristics, hemodynamics, and 1-year survival of patients with SLE-PAH (n = 110) with those of patients with SSc-PAH (n = 399) and IPAH (n = 1196) in the Registry to Evaluate Early and Long-term Pulmonary Arterial Hypertension Disease Management (REVEAL registry). They showed that patients with SLE-PAH had better hemodynamics and 1-year survival (94% vs. 93%) than those with IPAH. In addition, patients with SLE-PAH had a significantly higher 1-year survival rate than those with SSc-PAH (94% vs. 82%; p = 0·0009). Our results showed that the long-term survival of patients with SLE-PAH was significantly better than patients with IPAH (5-year survival 81·2% vs. 61·7%, p < 0.001; 10-year survival 66·6% vs. 44·1%, p < 0.001). A Favourable trend in overall survival has also been observed in patients with SLE-PAH over the last decade, consistent with IPAH^4^ and SSc-PAH^7^.

Consistent with previous studies that emphasised the importance of goal-oriented treatment of PAH^12, 21^, we proved that reaching a low-risk profile of PAH, which was recommended as a treatment goal during follow-up by the 2022 ECS/ERS Guidelines^18^. was a protective factor for survival in SLE-PAH. Moreover, SLE-PAH is more complicated than other forms of PAH in terms of the underlying autoimmunity and inflammation at its onset and progression. Therefore, achieving the treatment goals for SLE should be emphasised. LLDAS is an attainable treatment target for SLE, based on the principle of an “acceptable” level of disease activity in patients treated with low-dose glucocorticoids and stable immunosuppressive treatments. Achieving LLDAS was proven to be associated with a lower frequency of SLE flares in a previous large cohort study as well as a decreased probability of organ damage, better quality of life, and reduced mortality^22^. For the first time, our study demonstrated that achieving LLDAS was markedly associated with reduced mortality risk in patients with SLE-PAH. A higher proportion of patients in cohort B achieved a low-risk profile of PAH and reached the LLDAS of SLE during follow-up, which partially explains the improvement in survival in the last decade. Therefore, a goal-oriented dual treat-to-target strategy is recommended for the management of SLE-PAH to achieve better long-term outcomes.

Timely and effective application of PAH-targeted therapy and immunosuppressive therapy contribute to the achievement of treatment goals. Upfront dual PAH-targeted therapy was recommended for the initial treatment of PAH based on the improvements in patient outcomes observed in the AMBITION (A Study of First-Line Ambrisentan and Tadalafil Combination Therapy in Subjects with Pulmonary Arterial Hypertension) trial^23^. Upfront dual PAH-targeted therapy was favourably associated with a low-risk state of PAH in our study. Besides PAH-targeted medication, our study also showed that more patients were able to achieve the PAH treatment goal when treated with IIT as part of the baseline treatment. Previous studies have demonstrated the important role of autoimmune processes in PAH development, especially in the early phase before right heart reconstruction^6^. Several retrospective studies have examined the use of immunosuppressive agents in CTD-PAH, suggesting that immunosuppressants are associated with improvements in exercise capacity, hemodynamics, and long-term survival^6^. Responders to immunosuppressive therapy were mostly in the early stage of PAH^24^. Our results indicate that early initiation of IIT may reverse PAH progression and help achieve a low-risk state. Thus, IIT should be considered to have the same importance as PAH-targeted therapy in the management of patients with SLE-PAH. Apart from traditional immunosuppressive drugs, biological agents, including rituximab, belimumab, telitacicept, tocilizumab, and JAK inhibitors, for which indications are increasing in patients with SLE with different organ involvement, may also have potential therapeutic benefits for SLE-PAH^6^. A randomised clinical trial of rituximab demonstrated that patients with SSc-PAH and elevated cytokine levels showed improvement in both the 6-MWD and PVR at 24 weeks^25, 26^. Therefore, we believe that it is especially worthwhile to explore the treatment effects in patients with SLE-PAH, considering that this condition has more prominent inflammation than SSc-PAH.

Early detection is also essential to achieve treatment goals. In the present study, patients in cohort B had better clinical and hemodynamic states, indicating that more patients were diagnosed and treated at an early stage of PAH. Interestingly, a recent multi-center cohort study conducted in China that recruited patients from cardiovascular centers showed that the overall survival of patients with CTD-PAH was comparable to that of patients with IPAH. Although SSc-PAH accounted for a very small proportion (7·1%) of this cohort, the 5-year survival rate was still much worse than that of patients in the present study (approximately 70% vs. 81·2%)^3^. This may be partly related to the fact that patients referred to cardiovascular centers primarily had symptomatic PAH. Since PAH develops occultly in patients with SLE in most cases, clinical factors and serological biomarkers are studied to identify SLE patients at risk and allow early detection of PAH. Previous studies have suggested that Raynaud’s phenomenon, serositis, hyperuricaemia, anti-RNP, anti-SSA/SSB, and antiphospholipid antibodies are risk factors for SLE-PAH^27–29^. Recently, a risk-stratification model incorporating routine clinical parameters to predict the risk of PAH in patients with SLE was developed^29^. Genetic studies have shown that HLA-DQA1*03:02 is a genetic risk of SLE-PAH^30^. Patients with the above risk factors or those deemed to be high risk by the algorithm were recommended annual TTE screening to detect signs of pulmonary hypertension.

This study has some limitations. Firstly, the long-term survival of patients with high-risk group has a tendency of improvement, however, without statistical significance, which could be partly explain by the limited number of patients in this group. Future study is needed focusing the high-risk group of patients and studying the prognostic value of dual treat-to-target strategy. Secondly, for the nature of a real-world study, the prognostic value of treatment was evaluated by using risk-matching analysis. To further study the treatment strategy of SLE-PAH, randomized clinical trials are needed. Finally, as disease characteristics and prognosis of SLE-PAH may vary among different populations, we called for global cohort study of SLE-PAH to expand the research populations and further explore the possible racial difference of this disease.

Taken together, this is the largest prognostic study of SLE-PAH, based on the CSTAR-PAH cohort. Our findings, for the first time, illustrated the progress made in the early detection and management of patients with SLE-PAH over the last decade, which led to significant improvements in survival. We emphasised that meeting the treatment goals of both SLE and PAH were independently associated with reduced mortality, which support the dual treat-to-target strategy in SLE-PAH. Iinitial use of IIT, together with PAH-targeted combination therapy contributed to the early achievement of PAH treatment goals, which are recommended to implement in the real clinical practice of SLE-PAH.

## Data Availability

Data collected for the CSTAR-PAH cohort, including de-identified individual participant data and a data dictionary defining each field in the set, will be available to researchers who will provide a methodologically sound and ethically approved proposal within 24 months after the publication of this article. Data sharing requests should be submitted to the corresponding authors.

## Acknowledgements

The authors thank all investigators, study coordinators, and patients who participated in the study. Jiuliang Zhao, Qian Wang, Mengtao Li, and Xiaofeng Zeng conceived and designed the study. Xingbei Dong, Junyan Qian, Wei Wei, Miaojia Zhang, Xiao Zhang, Xiaofei Shi, Yisha Li, Xiaoping Hong, Qiang Shu, Shuhong Chi, Xin Dong, Ping Zhu, Rong Zhang, Zhuoli Zhang, Hongfeng Zhang, Xinwang Duan, Jing Xue, Shuhong Zhou, Hongbin Li, Dan Chen, Junwei Zhang, Zhuang Tian, Yongtai Liu collected patient data. Xingbei Dong, Junyan Qian, Jiuliang Zhao and Yanhong Wang analyzed data. All authors interpreted the results. Xingbei Dong, Jiuliang Zhao, Junyan Qian, Qian Wang, and Mengtao Li wrote the manuscript. All authors critically reviewed and revised the manuscript and approved the final version for publication. All authors had full access to the study data and final responsibility for the decision to submit for publication.

